# TTI-0102: A Novel Natural Controlled-Release Cysteamine Prodrug for Mitochondrial Disease and Cystinosis

**DOI:** 10.64898/2026.03.26.26347968

**Authors:** P. Rioux

## Abstract

**Background:** Cysteamine is the only disease-modifying therapy for nephropathic cystinosis and has shown promise in mitochondrial disorders, but its clinical utility is limited by poor tolerability due to high peak concentrations with existing formulations. TTI-0102 is a novel natural controlled-release cysteamine prodrug designed to provide sustained cysteamine exposure with improved tolerability.

**Methods:** A multi-center, randomized, single-blind, placebo-controlled Phase 2 trial enrolled 9 patients with MELAS syndrome caused by mtDNA m.3243A>G mutation (>50% heteroplasmy) and moderate disease severity (NMDAS score 15-45). Patients received placebo (n=3) or TTI-0102 at 2.75 g/day for one week then 5.5 g/day (n=6, equivalent to 2.5 g/day cysteamine base). Pharmacokinetic parameters, safety, and pharmacodynamic biomarkers including pyruvate, taurine, pantothenic acid, tryptophan, GSH/GSSG, lactate, GDF-15, and FGF-21 were assessed. Clinical efficacy was evaluated using the Modified Fatigue Impact Scale (MFIS) and 12-minute walk test.

**Results:** TTI-0102 demonstrated expected gastrointestinal side effects (nausea, vomiting, diarrhea) consistent with the cysteamine class, with dropout occurring in patients 50 kg receiving fixed 5.5 g/day dosing. Weight-based dosing at 60 ± 5 mg/kg TTI-0102 (~26 mg/kg cysteamine base equivalent) achieved sustained 24-hour cysteamine exposure with half the daily dose and peak concentrations lower than expected by dose proportionality, compared to approved formulations (Procysbi^®^: 56 mg/kg, peak 2.5 mg/L vs. TTI-0102: 26 mg/kg, peak ~2 mg/L). TTI-0102 significantly elevated pantothenic acid (plateauing at 2 weeks) and taurine levels, providing mitochondrial cofactor support and antioxidant effects. Statistically significant pharmacodynamic effects included increased plasma pyruvate (p=0.03) without lactate elevation, suggesting enhanced glycolytic flux, and decreased tryptophan (p<0.01), potentially reducing oxidative stress from neurotoxic kynurenine pathway metabolites. Interestingly, increase in plasma pyruvate and decrease in tryptophan were negligible at doses up to 40 mg/kg/day, optimal at 60 mg/kg/day, and slightly less at 65 mg/kg/day. GSH/GSSG measurements were confounded by sample stability issues. GDF-15, FGF-21, and 12-minute walk distance showed no treatment-related changes. Most notably, MFIS total scores demonstrated significant improvement in TTI-0102-treated patients at 60 mg/kg/day average dose compared to placebo (p=0.04). Polynomial regression revealed therapeutic onset at ~4 weeks, maximal benefit at ~12 weeks, and subsequent plateau.

**Conclusions:** This Phase 2 trial provides proof-of-concept that TTI-0102 is safe and well-tolerated in MELAS patients while treated with less than 65 mg/kg/day, with efficacy signals in fatigue reduction, a cardinal symptom affecting 71-100% of mitochondrial disease patients. The drug’s tri-faceted mechanism through sustained cysteamine, taurine, and pantothenic acid delivery addresses oxidative stress, mitochondrial energy metabolism, and cofactor deficiency. Significant MFIS improvement coupled with favorable modulation of pyruvate and tryptophan supports advancing TTI-0102 to larger Phase 2b/3 trials in mitochondrial disease employing weight-based dosing (60 ± 5 mg/kg), validated patient-reported outcomes, and minimum 12-week treatment duration. The same mechanism of cysteamine/cystine thiol-disulfide exchange in lysosomes that may benefit mitochondrial diseases also supports cystinosis treatment. An investigator-initiated study in cystinosis will evaluate whether once-daily TTI-0102 at 60 ± 5 mg/kg can maintain therapeutic WBC cystine levels, potentially offering improved adherence and quality of life compared to current twice-daily or four-times-daily regimens, and this weight-adjusted dosing strategy and pharmacodynamic biomarkers identified in the MELAS study are going to be used to inform the design of the planned Phase 2 study in Leigh syndrome, another mitochondrial disorder, in collaboration with the Children’s Hospital of Philadelphia (CHOP), with particular attention to dose optimization and biomarker-based assessment of pharmacological activity.

## Introduction

Cysteamine: Historical Context and Unmet Need

Cysteamine occurs naturally in humans at very low levels. Immediate-release cysteamine (first hydrochloride, then bitartrate) was initially used in the USSR for radiation poisoning treatment in the 1950s and subsequently in the United States. Since 1987, cysteamine has been used for cystinosis treatment, receiving FDA approval in 1994.[1][2][3][4][5]

In cystinosis, white blood cell (WBC) cystine levels serve as the primary therapeutic target for monitoring cysteamine treatment. A strict 6-hour dosing interval of high-dose Cystagon^®^ is mandatory to maintain WBC cystine <1 nmol ½ cystine/mg protein, requiring up to 4 g/day for adult patients.[2][6] This regimen produces significant side effects (nausea, vomiting, bad odor, bad breath) around the time of high peak concentrations every 6 hours, necessary to maintain continuous minimal cysteamine concentration of ~2.57 µM (0.2 mg/L).[2]. The final dose is a compromise resulting in a trial-and-error process to control WBC cystine and tolerability. In the FDA Cystagon^®^ package insert there is a table for determination of maintenance dose in ranges of mg/kg vs. weight, which corresponds to a unique linear dose vs weight relation without ever mentioning this relationship as such.

A delayed-release formulation, Procysbi^®^, was approved in 2013 with every 12-hour dosing instead of 6-hour dosing, but with the same total daily dose and similar pharmacokinetics, resulting in even higher peak concentrations and side effects.[7][8]

The clinical significance of achieving target WBC cystine levels is substantial. A large international cohort study of 453 patients demonstrated that higher mean leukocyte cystine levels were a significant risk factor for early progression to end-stage kidney disease, while lower levels were associated with improved linear growth.[9] Clinical trial data showed that patients achieving good cystine depletion (median levels <1 nmol ½ cystine/mg protein) had creatinine clearance 20.8 mL/min/1.73 m^2^ greater than those with poor depletion (median levels >3 nmol ½ cystine/mg protein), despite being older.[2] Early cysteamine initiation (5 years of age) is associated with delayed kidney failure and reduced incidence of diabetes, hypothyroidism, and neuromuscular complications.[4][6]

The strict dosing schedule for cysteamine—every 6 hours for Cystagon® (immediate-release) or every 12 hours for Procysbi® (delayed-release)—is dictated by two fundamental pharmacological principles. First, cysteamine has a short elimination half-life (approximately 90 minutes for immediate-release and 253 minutes for delayed-release formulations), necessitating frequent dosing to maintain therapeutic levels. Second, cysteamine’s mechanism of action relies on thiol-disulfide exchange, a reversible chemical reaction that converts lysosomal cystine into cysteine and a cysteine-cysteamine mixed disulfide, both of which can exit the lysosome. This reaction is concentration-dependent and reversible—when cysteamine levels fall below a critical threshold, the reaction reverses and cystine reaccumulates.

There is no remanent effect. The strict adherence to dosing intervals is essential because the thiol-disulfide exchange mechanism requires sustained continuous minimal cysteamine concentrations to continuously remove accumulated cystine from lysosomes and prevent disease progression.

Since the 1980s, cysteamine has been successfully tested in multiple animal models for numerous indications beyond cystinosis: Huntington’s disease, Parkinson’s disease, non-alcoholic steatohepatitis (NASH), and mitochondrial diseases.[10][11][12] However, cysteamine has not progressed successfully in humans for these indications, where low doses have always been administered orally to prevent side effects due to high peak concentrations, and only twice-daily for compliance.[12]

There is clearly an unmet need for a tolerable cysteamine preparation in patients with cystinosis, and such preparation has the potential for benefiting patients with a variety of other diseases.

### TTI-0102: A Novel Cysteamine Prodrug

Thiogenesis Therapeutics has developed TTI-0102, a patented new chemical entity (NCE) and cysteamine prodrug. TTI-0102 is the acetate salt of the asymmetric disulfide formed by joining cysteamine and pantetheine, designed to yield two cysteamine molecules upon chemical and enzymatic degradation in the gastrointestinal (GI) tract.

The first step in TTI-0102 degradation is disulfide bond reduction, yielding cysteamine and pantetheine. This step occurs principally in the small intestine. The pantetheine is not absorbed; it is hydrolyzed over a period of hours by the enzyme pantetheinase in the cells of the GI mucosa to produce a second molecule of cysteamine, along with a molecule of pantothenic acid (vitamin B5—a useful marker of cysteamine produced from pantetheine). Neither TTI-0102 nor pantetheine is absorbed; they are converted into cysteamine and pantothenic acid in the GI tract, and the cysteamine is absorbed across the entire length of the GI tract, including the colon.

Three dose levels of TTI-0102 (600 mg, 1,200 mg, and 2,400 mg cysteamine base equivalent) were first tested in 80+ kg healthy volunteers, in comparison to 600 mg of immediate-release cysteamine (Cystagon^®^). This showed that contrary to the strict every 6-hour dosing of 600 mg of Cystagon® needed for maintaining efficacy (as measured by white blood cell cystine level in the treatment of adults with cystinosis), 5.5 g of TTI-0102 (i.e., 2,400 mg of cysteamine base equivalent) had the potential for once-daily dosing with no or minimal side effects (slight skin odor).

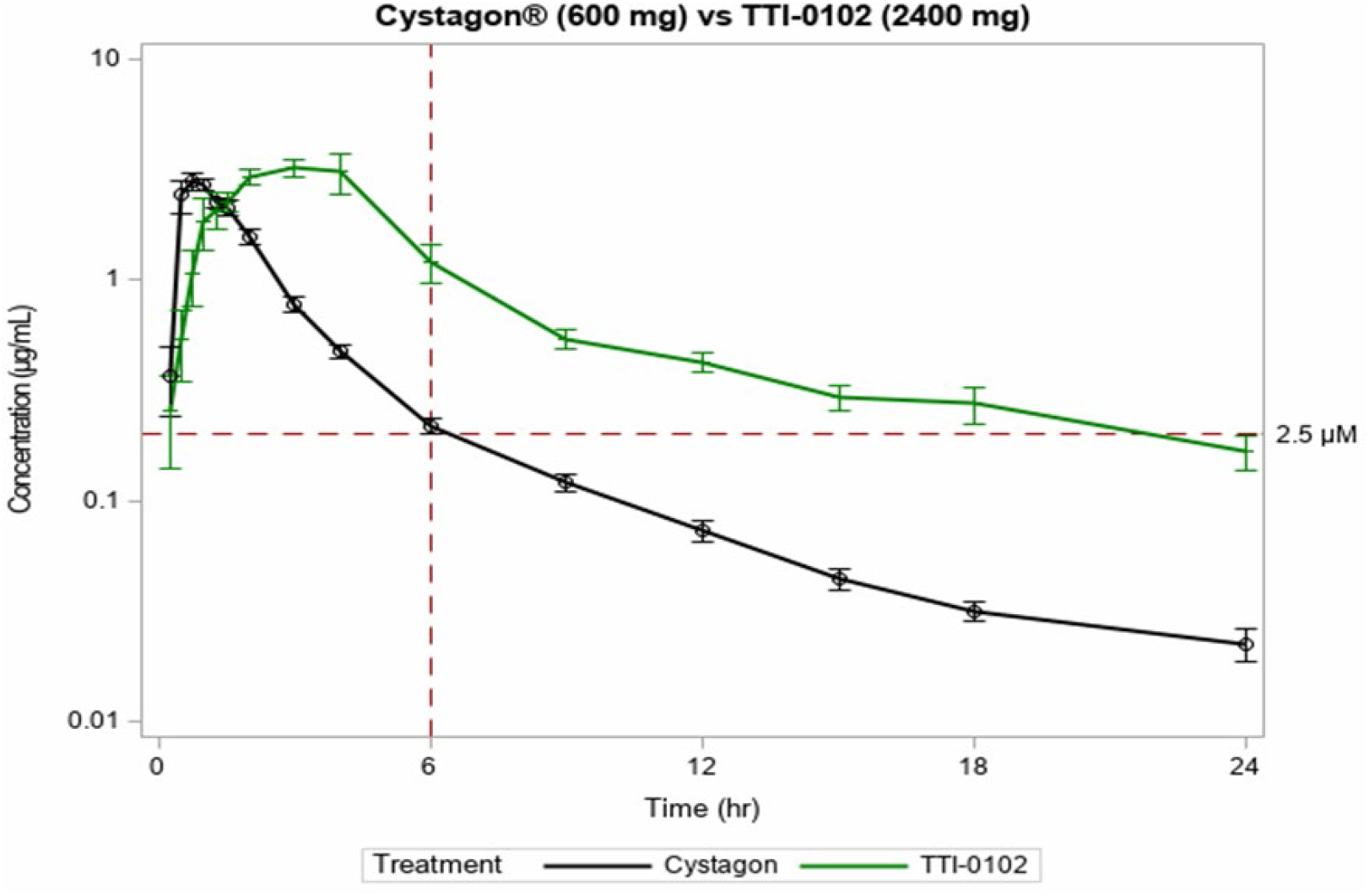

The goals of the MELAS study were to determine whether once-daily dosing was feasible, identify a dose that could minimize peak-related side effects, and establish biomarkers that could be used to predict efficacy. Moreover, this study explored whether cysteamine could be effectively utilized for conditions beyond cystinosis.

### MELAS Study Design

A multi-center, randomized, single-blind, placebo-controlled study was conducted to assess the efficacy, safety, tolerability, pharmacokinetics, and pharmacodynamics of oral TTI-0102 for treatment of patients with mitochondrial encephalomyopathy, lactic acidosis, and stroke-like episodes (MELAS).

Nine patients (39 to 82 kg) were enrolled, with 3 patients receiving placebo and 6 patients receiving 2.75 g once-daily dosing of TTI-0102 for one week, then 5.5 g daily dosing (i.e., 2.5 g/day of cysteamine base equivalent).

To be included, patients (males and females between 16 and 60 years of age) must have been diagnosed with MELAS, defined as:

- mtDNA mutation known to be associated with MELAS and MELAS phenotype, including but not limited to: m.3243A>G, m.13513G>A, m.10191T>C, m.3271T>C, m.13136_15374del, m.8363G>A. Mutation must have heteroplasmy >50% characterized by mutation load in urinary epithelium or blood. AND
- Two or more of the following clinical symptoms indicative of MELAS phenotype: diabetes, myopathy, seizures, at least one historic stroke-like episode, and exercise intolerance.
- With moderate disease severity defined as Newcastle Mitochondrial Disease Adult Scale (NMDAS) score between 15 to 45 inclusive.

### Safety

There were no unexpected side effects. Only expected side effects (as listed in the Cystagon® and Procysbi® label/package insert for the treatment of patients with cystinosis) were observed in MELAS patients treated with TTI-0102: nausea, vomiting, diarrhea.[2][8] These side effects are linked to the peak plasma concentration of cysteamine, as seen after administering high doses of Cystagon® and Procysbi® necessary to effectively treat patients with cystinosis.

In previous studies in patients with diseases other than cystinosis (neurodegenerative diseases, mitochondrial diseases, or NASH), these side effects were less frequent and milder because patients were administered lower doses than those used for cystinosis. However, it appeared that these trials could have been positive with higher doses of cysteamine, similar to the high doses (in mg/kg) used for the treatment of cystinosis—i.e., up to 4 g of cysteamine base equivalent per day for a 70 kg adult (with dose adjusted in mg/kg to the weight of the patient) to provide continuous minimal exposure to cysteamine.[10][11][12]

Oral administration of a single dose of 5.5 g (2.4 g of cysteamine base equivalent) of TTI-0102 to healthy volunteer adults has shown that it is possible to provide the same continuous minimal exposure to cysteamine over 24 hours with none or very few side effects after administering this single dose of 60 ± 5 mg/kg of TTI-0102 (~26 mg/kg of cysteamine base equivalent).

In the present trial, this fixed dose of 5.5 g/day was administered to all adult patients, but those weighing less than 50 kg (down to 39 kg or 43 kg) had significant side effects and dropped out.

Once PK data was available, study recruitment was terminated (closing the study after 9 rather than 12 participants were enrolled). During a planned interim review, PK data informed us that plasma cysteamine concentration was very high in low body weight participants, resulting in well-known and expected adverse events related to cysteamine, preventing subjects from wanting to complete the entire treatment period.

### Pharmacokinetics

#### Cysteamine Concentration vs. Dose

In the concentration-dose analysis, each datapoint corresponds to the plasma concentration of each patient (ID), either at C_max_ at 3 hours post-dose or at C_trough_ 24 hours post-dose. In green are the concentrations after 2.75 g dose during the first week, and in red the concentrations after 5.5 g dose after the first week.

Taking into account the side effects listed above and the corresponding plasma cysteamine concentrations at C_max_ and C_trough_, these results confirm that a single daily dose of 60 mg/kg of TTI-0102 (~26 mg/kg of cysteamine base equivalent) provides continuous minimal exposure to cysteamine with minimal side effects.

In comparison with Procysbi^®^, FDA and EMA approved for the treatment of cystinosis, to achieve minimal exposure over 24 hours, the total daily dose of cysteamine base equivalent of Procysbi^®^ was 56 mg/kg which, given twice a day, led to an average peak concentration of 2.5 mg/L.[7][8] With TTI-0102, we need half the dose (26 mg/kg of cysteamine base equivalent), given only once a day, which would lead to a lower peak (~2 mg/L), meaning fewer side effects.

#### Pantothenic Acid (Vitamin B5) Concentration vs. Time

Oral administration of one molecule of TTI-0102 results in absorption of one molecule of pantothenic acid and two molecules of cysteamine. After ~2 weeks, the plasma concentration of vitamin B5 plateaus. Pantothenic acid is absorbed efficiently from the gastrointestinal tract and distributed throughout the body, with high concentrations in the liver and colonic mucosa. Direct human pharmacokinetic data on half-life is limited. Animal studies provide more detail: in mice, the half-life of pantothenic acid in the liver is approximately 69–82 hours, and in the brain 136–144 hours, following oral administration. These values reflect tissue turnover rather than plasma half-life but suggest pantothenic acid and its metabolites have a tissue half-life of 2–6 days.

Pantothenic acid is required for the biosynthesis of coenzyme A (CoA), which is central to mitochondrial energy metabolism, including the tricarboxylic acid (TCA) cycle and fatty acid oxidation.[13][14][15] Deficiency in pantothenic acid impairs mitochondrial enzymes and energy production, as seen in neurodegenerative diseases like Alzheimer’s, where reduced pantothenate leads to decreased activity of CoA-dependent TCA cycle enzymes.

Pantothenic acid and its derivatives protect mitochondria from oxidative damage by increasing cellular glutathione levels, a major antioxidant. This effect is mediated by boosting ATP production, which in turn supports glutathione biosynthesis.[16] Pantothenic acid does not act as a direct free radical scavenger but rather enhances the cell’s endogenous antioxidant capacity, mainly through glutathione-dependent pathways. This protection is particularly relevant under conditions of oxidative stress, where pantothenic acid or its derivatives (e.g., panthenol) restore mitochondrial redox balance and energy metabolism.

#### Taurine Concentration vs. Time

As expected, since taurine is the main metabolite of cysteamine, there is a significant increase of taurine, as measured at the time of Ctrough of cysteamine, after administration of TTI-0102 compared to its physiological level under placebo, with most of the time concentration under limit of quantification. Like for cysteamine concentration, higher doses in mg/kg of TTI-0102 produce higher residual concentrations of taurine.

Taurine is highly concentrated in tissues with high oxidative metabolism and localizes to mitochondria, where it acts as a pH buffer in the mitochondrial matrix, stabilizing the environment for oxidative phosphorylation and preventing excessive reactive oxygen species (ROS) production.[17][18][19][20] Taurine deficiency leads to mitochondrial dysfunction, increased ROS, and impaired electron transport chain activity, which can trigger apoptosis. Experimental studies show that taurine supplementation increases GSH levels and glutathione peroxidase activity, especially in aging or stressed tissues, further enhancing mitochondrial antioxidant capacity.[21]

The cytoprotective effect of taurine has attracted significant recent attention. Clinical studies in several degenerative diseases have yielded encouraging results, culminating in the approval of taurine in Japan for therapy of congestive heart failure and MELAS. However, according to the literature, it is difficult to generalize about the beneficial effect of exogenous antioxidants in preventing cardiovascular disease. This does not necessarily seem to be the case for endogenous antioxidants. As a matter of fact, in humans, plasma taurine levels rise sharply within 1–2 hours after an oral dose, peaking at approximately 10–13 times baseline, and then decline over several hours, approaching baseline by 4–6 hours post-dose. Like for cysteamine or other thiols, constant minimal exposure over 24 hours, as provided by administration of TTI-0102, seems critical for efficacy.

### Pharmacodynamics

#### GSH and GSSG

Cysteamine, by interacting with cystine in the lysosome (where cystine is stored in patients with cystinosis but also in normal subjects), leads to production of cysteine in the cytoplasm. Cysteine is used to build glutathione, the only small antioxidant that can be transported into mitochondria, where it is used to neutralize reactive oxygen species (ROS).[6][22][23]

The thiol-disulfide exchange reaction to generate cysteine is not limited to cysteamine but has been characterized with another small thiol, N-acetylcysteine (NAC). As a matter of fact, it appears that in vivo, the concentration of NAC is not high enough for NAC to be converted into cysteine, but NAC interacts with cystine to generate free cysteine and a mixed disulfide, NAC-cysteine. This free cysteine is then used to enhance cellular glutathione levels, but the concentration of NAC for this reaction to happen needs to be far higher than with cysteamine. N-acetylcysteine (NAC), 2-mercaptoethanesulfonic acid (MESNA), and mercaptopropionylglycine (MPG) elevate cysteine levels to a lesser extent than cysteamine (2-fold as compared to 6-fold in the case of cysteamine).

Although in this study, this potential mechanism of action (MoA) for mitochondrial diseases is supported by PK data, with “dose × duration of exposure” relationship linked to patient’s weight (like for the treatment of cystinosis with cysteamine), PD data could not directly confirm the potential mechanism of action, since there was no difference in whole blood GSH and GSSG levels between placebo and TTI-0102.[2][6] It was very difficult to develop an ICH method for measuring these compounds, and analyses were performed on blood frozen for more than 6 months for most of the samples, which probably impacted the stability of these samples. Results were very different from what had been previously published for mitochondrial diseases, especially regarding GSSG concentrations. Only a very small percentage of GSH (5%) exists in its oxidized form GSSG under normal conditions. As a matter of fact, when exposed to oxygen in the air, GSH is rapidly oxidized into GSSG, which could explain why the concentration of GSSG in the samples was very high but not very different from equilibrium with GSH.

#### Lactate and Pyruvate

There was no difference in plasma concentration of lactate between treatment and placebo over time, but TTI-0102 oral administration led to significant increase in pyruvate, which in this case of stable lactate may indicate enhanced glycolytic flux and potential for increased ATP production.

Cysteamine can boost the activity of thiol-containing enzymes such as pyruvate kinase, which is crucial for glycolysis and energy homeostasis. This might be the reason why oral administration of TTI-0102 has a statistically significant effect on pyruvate concentration, despite the small number of subjects (p=0.03, as determined by repeated measure mixed model ANOVA with baseline and AR(1) covariance matrix).

Pyruvate and glutathione are metabolically and functionally linked through several mechanisms involving antioxidant defense, redox balance, and biosynthesis.[24][25][26][27][28][29]

Pyruvate is the end product of glycolysis and is transported into mitochondria via the mitochondrial pyruvate carrier (MPC). Once inside, pyruvate is converted by the pyruvate dehydrogenase complex (PDH) to acetyl-CoA, entering the Krebs cycle to support ATP production and biosynthesis of key metabolites. Pyruvate can also directly scavenge H_2_O_2_, providing additional protection against oxidative stress, but this protective effect is dependent on a functional mitochondrial glutathione pool.[25]

Pyruvate serves as a key metabolic intermediate that can enhance glutathione synthesis and maintain cellular antioxidant capacity. Specifically, pyruvate carboxylase activity is required for glutathione synthesis, as it channels glucose-derived carbon into pathways that support de novo glutathione production and help restrict oxidative stress, particularly in pancreatic islets. This metabolic axis is critical for cellular survival under stress conditions.

Pyruvate also supports the glutathione redox cycle by improving NADPH supply, which is necessary for the regeneration of reduced glutathione (GSH) from its oxidized form (GSSG). In endothelial cells exposed to high glucose, pyruvate supplementation increases glucose oxidation through the pentose phosphate pathway, boosts NADPH levels, and enhances glutathione-dependent degradation of hydrogen peroxide, thereby protecting against oxidative injury.

The observed increase in mitochondrial pyruvate sensitivity during acidosis may reflect an adaptive mechanism in muscle tissue. By prioritizing carbohydrate-derived substrates over fatty acid-derived substrates under high-intensity conditions, muscle cells may optimize ATP production by shifting towards the more rapidly metabolized energy source. This shift would also enhance post-exertion recovery once oxygen availability is restored.

Additionally, pyruvate can induce glial cells to upregulate glutathione synthesis, providing neuroprotection against excitotoxicity. In hepatocytes and renal cells, pyruvate prevents glutathione depletion and supports redox homeostasis, which is essential for cell viability and protection against various toxic insults.

Conversely, glutathione is necessary for the protective effects of pyruvate; in its absence, pyruvate can exacerbate oxidative damage, particularly in mitochondria, due to increased reactive oxygen species production.[25]

In summary, pyruvate promotes glutathione synthesis and function, while glutathione is essential for pyruvate’s protective antioxidant effects, forming a reciprocal relationship crucial for cellular redox balance and defense against oxidative stress.

#### Tryptophan

Tryptophan (as included in full amino acid panel for each subject) is primarily metabolized via the kynurenine pathway, producing several catabolites (TRYCATs) that can directly affect mitochondrial function. Notably, some TRYCATs such as quinolinic acid are neurotoxic and can inhibit mitochondrial ATP production, increase oxidative and nitrosative stress, and provoke mitochondrial damage. This mitochondrial dysfunction leads to increased production of reactive oxygen species (ROS).

Oral administration of TTI-0102 led to a statistically significant tryptophan decrease from baseline (p<0.01, as determined by repeated measure mixed model ANOVA with baseline and AR(1) covariance matrix).

Tryptophan loading in humans and animals has been shown to increase markers of oxidative stress, including lipid peroxidation and depletion of antioxidant defenses. Experimental data show that glutathione can prevent tryptophan-induced lipid peroxidation and oxidative damage in vitro and in vivo. Tryptophan metabolism supports ATP production by supplying NAD+ via the kynurenine pathway, thereby enabling ATP synthesis and mitochondrial energy generation.

#### GDF-15 FGF-21

No significant difference in growth differentiation factor 15 (GDF-15) and fibroblast growth factor 21 (FGF-21) changes from baseline were seen in these patients with MELAS over time. GDF-15 and FGF-21 are consistently elevated in patients with mitochondrial diseases, particularly those with muscle-manifesting mitochondrial translation defects or mtDNA deletions, and their levels correlate with disease severity. However, their utility as markers of treatment response is less well established.

GDF15 has shown promise as a marker of therapeutic response in TK2 deficiency: In patients with TK2-deficient myopathy treated with deoxynucleosides, GDF15 levels declined significantly in parallel with clinical improvement, suggesting its potential as a biomarker for monitoring treatment efficacy in this specific context. FGF21 levels also decreased, but less consistently and to a lesser extent. This supports the use of GDF15, and to a lesser degree FGF21, for tracking response to therapy in TK2 deficiency.

For other mitochondrial diseases, the evidence is insufficient to support GDF-15 or FGF-21 as general markers of treatment efficacy. Most studies focus on their diagnostic and prognostic value, with only limited longitudinal data on changes in biomarker levels in response to therapy. The literature suggests that while these biomarkers reflect disease activity and severity, their responsiveness to treatment outside TK2 deficiency remains to be validated.

### Clinical Evaluations

#### 12-Minute Walk Test

There was no change in maximum distance walked in 12 minutes by MELAS adult patients, treated or not treated, whatever the duration of treatment each completed.

The validity of the 12-minute walk test (12MWT) in mitochondrial disease clinical trials is not well-established, as the available evidence is limited primarily to McArdle disease (glycogen storage disease type V), which is a distinct metabolic myopathy rather than a primary mitochondrial disorder. The 6-minute walk test (6MWT) has been more extensively studied as an outcome measure, but endurance-based walking tests have limitations in this population.

#### Modified Fatigue Impact Scale (MFIS)

A new study reports that in MELAS, patient-reported outcomes don’t always match objective measures—highlighting the need for multi-dimensional outcomes in mitochondrial disease trials.

The Modified Fatigue Impact Scale (MFIS) total score is such a multi-dimensional outcome measure: it is used to quantify the perceived impact of fatigue on physical, cognitive, and psychosocial functioning in patients with mitochondrial disease, serving as a validated patient-reported outcome measure for both clinical assessment and research.[30][31][32] Scores are interpreted to reflect the degree to which fatigue interferes with activities, with higher scores indicating greater impact. For example, a score ≥40 suggests excessive symptomatic fatigue, while scores ≥80 indicate severe, functionally limiting fatigue.

The MFIS is particularly valuable because fatigue in mitochondrial disease is often under-recognized and not fully explained by clinical or genetic assessments alone.[31] Patient-reported outcomes like the MFIS provide critical information about quality of life and daily functioning that may not be captured by objective measures. The MFIS has demonstrated validity and reliability in chronic disease populations, including mitochondrial disorders, and is recommended for use in both clinical practice and research settings to monitor fatigue and guide individualized management.

In our study, MELAS patients treated with TTI-0102 expressed significant improvement compared to those treated with placebo. This change is statistically significant (p=0.04, as determined by repeated measure mixed model ANOVA with baseline and AR(1) covariance matrix), despite the limited number of patients.

Using polynomial regression for the graphic shows that it takes ~4 weeks before the start of beneficial effect on fatigue; this effect is maximum at ~12 weeks, and the subjects don’t feel any significant improvement afterwards.

Fatigue is a cardinal symptom in mitochondrial disorders, including MELAS. Studies demonstrate that 71-100% of patients with primary mitochondrial disease report fatigue, with 62% experiencing excessive symptomatic fatigue and 32% reporting severe, functionally limiting fatigue.[30][32] Fatigue severity correlates with disease severity and remains stable over time in the majority of patients.

## Discussion

This randomized, placebo-controlled trial in MELAS patients provides preliminary evidence supporting the therapeutic potential of TTI-0102, a novel controlled-release cysteamine prodrug designed to overcome tolerability challenges associated with traditional cysteamine therapy.

### Safety and Dosing Considerations

The trial demonstrated that weight-based dosing of TTI-0102, as well as strict adherence to daily dosing, is critical for optimizing the therapeutic window. Administration of a fixed 5.5 g/day dose (equivalent to 2.4 g cysteamine base) resulted in significant gastrointestinal side effects and patient dropout among individuals weighing less than 80 kg, as plasma cysteamine peak concentrations (Cmax) at 3 hours post-dose were substantially elevated in these patients. These findings align with preclinical studies demonstrating the narrow therapeutic window of cysteamine, with toxicity correlating with excessive hydrogen peroxide production at millimolar concentrations. The study confirms that a weight-adjusted dose of approximately 60 ± 5 mg/kg TTI-0102 (~26 mg/kg cysteamine base equivalent) provides sustained minimal cysteamine exposure over 24 hours with minimal adverse effects—representing half the dose required with Procysbi^®^, the approved delayed-release formulation for cystinosis, while achieving comparable trough levels and lower peak concentrations.[7][8]

Based on these observations, the weight-adjusted dosing strategy and pharmacodynamic biomarkers identified in the MELAS study have been used to inform the design of the planned Phase 2 study in Leigh syndrome, another mitochondrial disorder, in collaboration with the Children’s Hospital of Philadelphia (CHOP), with particular attention to dose optimization and biomarker-based assessment of pharmacological activity.

### Pharmacokinetic and Metabolic Insights

TTI-0102’s unique dual-release mechanism yielded pharmacologically relevant elevations in both pantothenic acid (vitamin B5) and taurine, metabolites with established roles in mitochondrial function.[13][14][15][16][17][18][19][20][21] Pantothenic acid, essential for coenzyme A biosynthesis and mitochondrial energy

This multicenter study was approved by:

**France EC**

Comité de protection des personnes (CPP) SUD Mediterranee V

**Netherlands EC**

Radboud University Medical Center - METC East Netherlands

## Data Availability

All data produced in the present study are available upon reasonable request to the authors

## Acknowledgement

We are very thankful to the patients and the clinical teams of Radboud University Nijmegen Medical Centre (Netherlands) and Centre Hospitalier Universitaire d’Angers (France) for their participation in this operationally challenging study.

